# Effects of *Lactiplantibacillus plantarum* KABP051 Probiotic on Body Composition, Microbiome and Mood in Healthy Overweight Adults

**DOI:** 10.1101/2025.08.01.25332799

**Authors:** Shawn Talbott, Bret Stephens, Julie Talbott, Marc Oddou, Aoki Fumiki

**Author notes:** Address correspondence to: Shawn Talbott, 3 Waves Wellness 73 Warren Avenue, Plymouth, MA 02360, United States of America.

## Abstract

Obesity and mental health disorders are among the greatest public health challenges of the twenty-first century. Interestingly, an altered microbiome profile has been associated with both conditions. The aim of this randomized, double-blind, placebo-controlled clinical trial was to evaluate the effects of dietary supplementation with a specific probiotic strain (*Lactiplantibacillus plantarum* KABP051) on body composition and gut microbiome balance, together with measures of mood state, in a population of healthy overweight subjects. Sixty healthy, moderately stressed, non-depressed and overweight or obese volunteers were supplemented for 12 weeks with probiotic (*L. plantarum* KABP051; 1 billion cfu/day) or placebo (microcrystalline cellulose). The KABP051 group experienced significantly greater improvements compared to placebo on body composition measurements, including a reduction in body weight and waist circumference, which decreased in 1.97±0.77 (mean±SE) kg and 2.15±0.81 (mean±SE) cm vs. placebo at the end of the intervention (both p< 0.05, MMRM and post-hoc analysis). Microbiome composition improved in KABP051 group, with significant increase in the relative abundance of *Lactiplantibacillus* spp. vs. placebo. Body fat percentage, POMS fatigue, and confusion sub-scores showed a global trend toward improvement compared to placebo, with the change at 12 weeks being significant in the three measurements in post-hoc analysis (p=0.015, p=0.014 and p=0.016, respectively). No serious adverse events were registered during the intervention period. These results suggest that a specific strain of probiotic bacteria (*L. plantarum* KABP051) may have both metabolic and psychobiotic effects and may be beneficial for enhancing weight loss and body composition, improving energy and mood levels while embarking on a healthy lifestyle regimen.

## INTRODUCTION

Globally, approximately 3 billion people struggle with the so-called twin epidemics of obesity and depression.^1–6^ Nearly two decades have passed since the initial discovery of the close link between the gut microbiota and obesity^6–7^ and a large and growing body of evidence is elucidating the underlying biological mechanisms whereby the gut microbiome may influence appetite, energy harvest from food, and fat storage/expenditure.^8–11^ One of the predominant mechanisms underlying the microbiome/obesity relationship is the metabolic endotoxemia hypothesis, whereby an impaired or permeable (leaky) gut barrier allows translocation of endotoxins from the gut lumen into systemic circulation, thereby leading to low-grade inflammation and metabolic disorders including obesity, diabetes, and metabolic syndrome.^12–14^ Similarly, the gut microbiome is also closely linked to psychological mood states, including depression, anxiety, stress, and fatigue through multiple communication pathways, including neurotransmitters, the immune system, and the inflammatory cytokine cascade via the gut-brain axis.^5,15–17^ A close bidirectional relationship between overweight and depression has repeatedly been established; thus, being overweight increases the risk of developing depression, and having depression increases the risk of becoming overweight.^15–17^ In addition, antidepressant medications often lead to weight gain,^17^ and dietary restriction for weight loss often exacerbates depression.^19^

Probiotics are live micro-organisms that confer health benefits to the host along the gut-brain axis^5,20^ with an emerging class of specialized bacterial strains termed psychobiotics that confer psychological benefits related to mood and cognition.^24^ A rapidly growing number of recent clinical trials have shown both promising weight management benefits of probiotics,^28–35^ as well as reductions in anxiety and depression levels with specific probiotic strains,^5,36–40^ while it has been acknowledged that probiotic effects are strain-specific.^20^

*Lactiplantibacillus plantarum* (*L. plantarum*) species can inhabit a wide variety of ecological niches including the human gastrointestinal mucosa. They possess genetic and functional characteristics that make them good candidates for ideal probiotics.^21^ *L. plantarum* KABP051 is a human-derived strain isolated from a South American individual. It has been shown to produce metabolites such as 2-hydroxyisocaproic acid (HICA) and 3-phenyllactic acid (PLA),^22^ which have antimicrobial properties and may offer additional health benefits.^23^

The present study aimed to determine the effects of this specific strain of probiotic bacteria *L. plantarum* KABP051 on body composition, microbiome composition and psychological parameters in a population of healthy, overweight and obese as well as moderately stressed adults.

## MATERIALS AND METHODS

### Study design

This was a randomized, placebo-controlled, double-blind study. The protocol was approved by the Institutional Review Board (IRB) and Human Subjects Committee (HSC) at Marietta College (Marietta, OH, USA; Protocol #09nct162022-2) and registered at ClinicalTrials.gov (NCT06808061). This study was conducted following the guidelines laid down in the Declaration of Helsinki in 2013.

Sixty healthy overweight and moderately stressed volunteers were recruited to participate in a research study investigating the effects of a specific probiotic strain (*Lactiplantibacillus plantarum* KABP051) on measurements of microbiome balance; body composition (weight, waist circumference, fat mass); and psychological mood state. The main inclusion criteria included age between 18 and 65 years; BMI range of 25-35; body fat percentage of 20-30%, or waist circumference of 25-30 inches (63.5-76.2 cm); and moderate stress on the screening survey based on the Perceived Stress Scale PSS-10. The main exclusion criteria included: (1) pregnant women; (2) current use of incompatible medications or supplements (such as other probiotics and supplements or medication for weight loss, stress/depression, blood sugar). Volunteers who met the inclusion/exclusion criteria completed and signed an informed consent form and were enrolled in the trial and randomly allocated to receive either probiotic (*L. plantarum* KABP051; 1 billion colony forming units [cfu]) or matching placebo (PL; microcrystalline cellulose) for a period of 90 days, according to a computer-generated randomization list. *L. plantarum* KABP051 and placebo capsules were supplied by Kaneka Probiotics (Kaneka Americas Holding, Inc.), were indistinguishable and were delivered in coded, anonymous boxes. In addition, the probiotic capsules were monitored to confirm to meet the criteria of >1 billion cfu/capsule throughout the study period.

All participants were instructed to follow the same daily protocol to take one capsule of the assigned supplement or placebo daily during a meal (breakfast, lunch or dinner) with a beverage of their choice. Product compliance was evaluated by subject confirmation of supplement consumption and by measuring unused product at study completion.

Subjects also participated in weekly (via online webinar) lifestyle education sessions on nutrition, exercise, sleep, stress, and related healthy lifestyle topics, including a prudent diet/exercise/sleep regimen, but did not diet or workout per se. This periodic contact is considered extremely effective for maintaining compliance with the supplementation regimen and reducing dropouts.

### Study endpoints

Main and secondary study outcomes were measured at baseline and monthly at week 4, week 8, and week 12.

### Body composition and waist circumference (bioelectrical impedance)

The primary study endpoint was the evaluation of the efficacy of probiotic intervention on body weight, waist circumference and body fat. Body composition was evaluated by using an established DSM-BIA device (InBody 770, InBody USA, Cerritos, CA) to collect body weight and body fat percentage measurements. Previous validation studies of the accuracy of BIA technique using DEXA as reference standards have shown direct segmental multi-frequency BIA (DSM-BIA) to be superior in the estimation of body composition, with 99% accuracy compared to DEXA.^45^

### Mood assessment (POMS; Profile of Mood States)

Changes in mood, focus, and energy were assessed using the research-validated Profile of Mood States (POMS) questionnaire to measure six primary psychological factors (tension, mood, irritability, fatigue, confusion, and vigor) plus the combined global mood state (called total mood disturbance, TMD) as an indication of overall subjective well-being. TMD is calculated as (tension + depression + anger + fatigue + confusion) – vigor.^46^ The output of the POMS questionnaire is an assessment of the positive and negative moods of each healthy participant at baseline and post-supplementation.

### Microbiome assessment

Microbiome PCR analysis of fecal samples was performed using the complete BiomeTracker system (Wasatch Scientific, Murray, UT). Briefly, fecal samples were obtained using nylon swabs and placed into a preservative binding buffer to lock the composition of bacteria in place. DNA was purified using DNA columns, and ∼20 ng of DNA from each sample was added to the reaction mixtures. Samples were processed in duplicate using an ABI 7500 Fast (Applied Biosystems) instrument. Specific primers were used to measure total *Bacteroidetes*, total *Firmicutes*, total *Bifidobacterium*, total *Lactobacillus*, and total *Akkermansia*. Participants were provided with an at-home fecal microbiome collection kit with instructions to collect their sample. Samples were delivered in a pre-addressed/prepaid transport envelope to the laboratory for analysis.

### Salivary cortisol

For salivary cortisol analysis, participants were provided with an at-home saliva collection kit with instructions to collect a first-morning saliva sample before eating or brushing their teeth. Samples were delivered in a pre-addressed/pre-paid transport envelope to the laboratory for analysis. Cortisol in saliva diluted ≥1:4 per manufacturer’s recommendation was analyzed using an ELISA kit (Enzo Biochem, Inc., USA).

### Data management and analysis

Data were recorded on case report forms (CRFs) and typed into Excel worksheets. Participants were analyzed per allocated group, using all available data at each time point (full analysis set). Data were analyzed using Mixed Repeated Measures Models (MRMMs), with the allocated group and study visit as factors, and baseline values as covariates. Adjusted post-hoc analyses were performed for each time point within the MRMMs. For ponderal data, subject height was also included as a covariate, and microbiome data were log-transformed for analysis. Results are reported as adjusted mean changes from baseline and their standard errors. Data were analyzed using SPSS version 20.0 (SPSS Inc., Chicago, IL, USA). Significance was set at two-sided alpha<0.05.

## RESULTS

Baseline characteristics were comparable between study groups. Out of the 60 recruited subjects, 5 were early terminated due to loss of follow-up (2 in probiotic group and 3 in placebo group). The compliance rate of the completed subjects (N=55) was virtually 100% in both the KABP051 and placebo groups, based on study product consumption. There were no significant differences between groups in terms of subject demographics or the parameters for the primary and secondary outcomes of this study (Table 1).

**Table 1.** Subject characteristics at baseline.

|  | **Placebo** | **Probiotic** |
| --- | --- | --- |
| **Female (n (%))** | 24 (80%) | 25 (83%) |
| **Male (n, (%))** | 6 (20%) | 5 (17%) |
| **Age (years)** | 50.0 ± 9.0 (range 33-71) | 51.6 ± 11.6 (range 29-73) |
| **Height (cm)** | 167.1 ± 1.7 | 166.0 ± 1.5 |
| **Weight (kg)** | 82.0 ± 3.5 | 82.6 ± 3.6 |
| **Waist perimeter (cm)** | 98.8 ± 2.9 | 97.7 ± 2.8 |
| **Body fat %** | 36.9 ± 1.7 | 36.0 ± 1.7 |
| **Total Mood Disturbance (TMD)** | 35.6 ± 6.2 | 34.2 ± 5.4 |
| **- Tension sub-score** | 11.8 ± 1.2 | 11.1 ± 1.1 |
| **- Depression sub-score** | 11.3 ± 1.9 | 8.8 ± 1.1 |
| **- Anger sub-score** | 8.2 ± 1.4 | 7.6 ± 1.2 |
| **- Fatigue sub-score** | 9.6 ± 1.2 | 10.6 ± 1.1 |
| **- Confusion sub-score** | 8.3 ± 0.9 | 8.2 ± 0.8 |
| **- Vigor sub-score** | 13.6 ± 1.4 | 13.0 ± 1.3 |

Values are presented as number and percent or as mean and standard error.

### Main outcomes

Probiotic supplementation resulted in significant global reduction in total body weight (p=0.022) and waist circumference (p=0.027) compared to placebo, while body fat also showed a global trend (p=0.063) over the 12 weeks of treatment. In post-hoc analyses per timepoint, change at 12 weeks was significant for all three parameters (p=0.015, p=0.011 and p=0.025, respectively), compared to placebo (Fig. 1) (see details in Supplementary Table S1).

**Figure 1.**
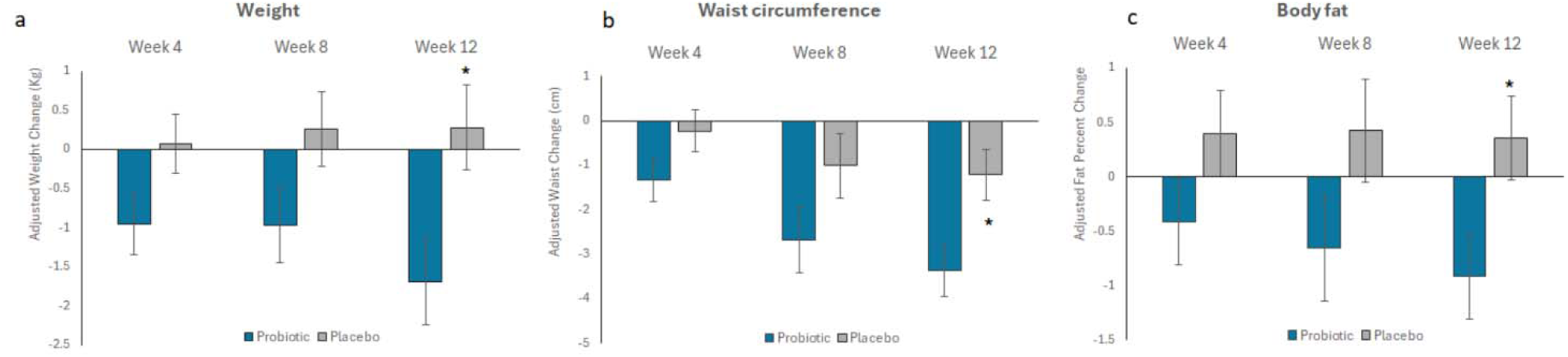
Changes from baseline in body weight (a), waist circumference (b) and percentage of body fat (c). Bars represent mean changes from baseline and their standard error. Asterisk (*) indicates significant difference vs. placebo (post-hoc analysis per timepoint from a mixed model for repeated measures, MMRM).

### Secondary outcomes

Regarding mood status (POMS), a global trend was observed in fatigue (p=0.096) and confusion sub-scores (p=0.095) compared to placebo, with significant change at 12 weeks for both parameters (p=0.014 and p=0.016, respectively) in post-hoc analysis (Table 2). Conversely, the other sub-scores and the overall TMD score did not show relevant changes. No serious adverse events were registered during the intervention period.

**Table 2.** Changes in POMS scores for fatigue, confusion, depression, and TMD outcomes versus baseline.

|  | Week 4 |  | Week 8 |  | Week 12 |  |
| --- | --- | --- | --- | --- | --- | --- |
|  | Placebo | Probiotic | Placebo | Probiotic | Placebo | Probiotic |
| **POMS scores** |  |  |  |  |  |  |
| **POMS fatigue** | -1.27<br>(±0.96) | -2.52<br>(±0.96) | -2.23<br>(±1.07) | -3.80<br>(±1.07) | -0.59<br>(±1.06) | -4.40<br>(±1.06)# |
| **POMS confusion** | -0.93<br>(±0.74) | -1.82<br>(±0.74) | -1.69<br>(±0.74) | -2.86<br>(±0.74) | -0.69<br>(±0.70) | -3.18<br>(±0.70)# |
| **POMS depression** | -0.64<br>(±1.93) | -2.63<br>(±1.93) | -1.72<br>(±1.41) | -4.83<br>(±1.41) | -1.88<br>(±1.10) | -5.39<br>(±1.10) |
| **POMS TMD** | -7.45<br>(±5.86) | -10.86<br>(±5.86) | -12.13<br>(±5.56) | -20.22<br>(±5.56) | -9.01<br>(±5.09) | -21.58<br>(±5.09) |

Data are expressed as mean change from baseline (± standard error);

Among the bacterial components of the microbiota, global significant changes were observed for *Lactobacillus* spp. only (p=0.048), the post-hoc analysis showing a significant effect at 12 weeks (p=0.047) (Fig. 2). No differences were found between KABP051 and PL in the *Bacteroidetes*, *Firmicutes*, *Bifidobacterium* spp., and *Akkermansia* spp. bacterial groups.

**Figure 2.**
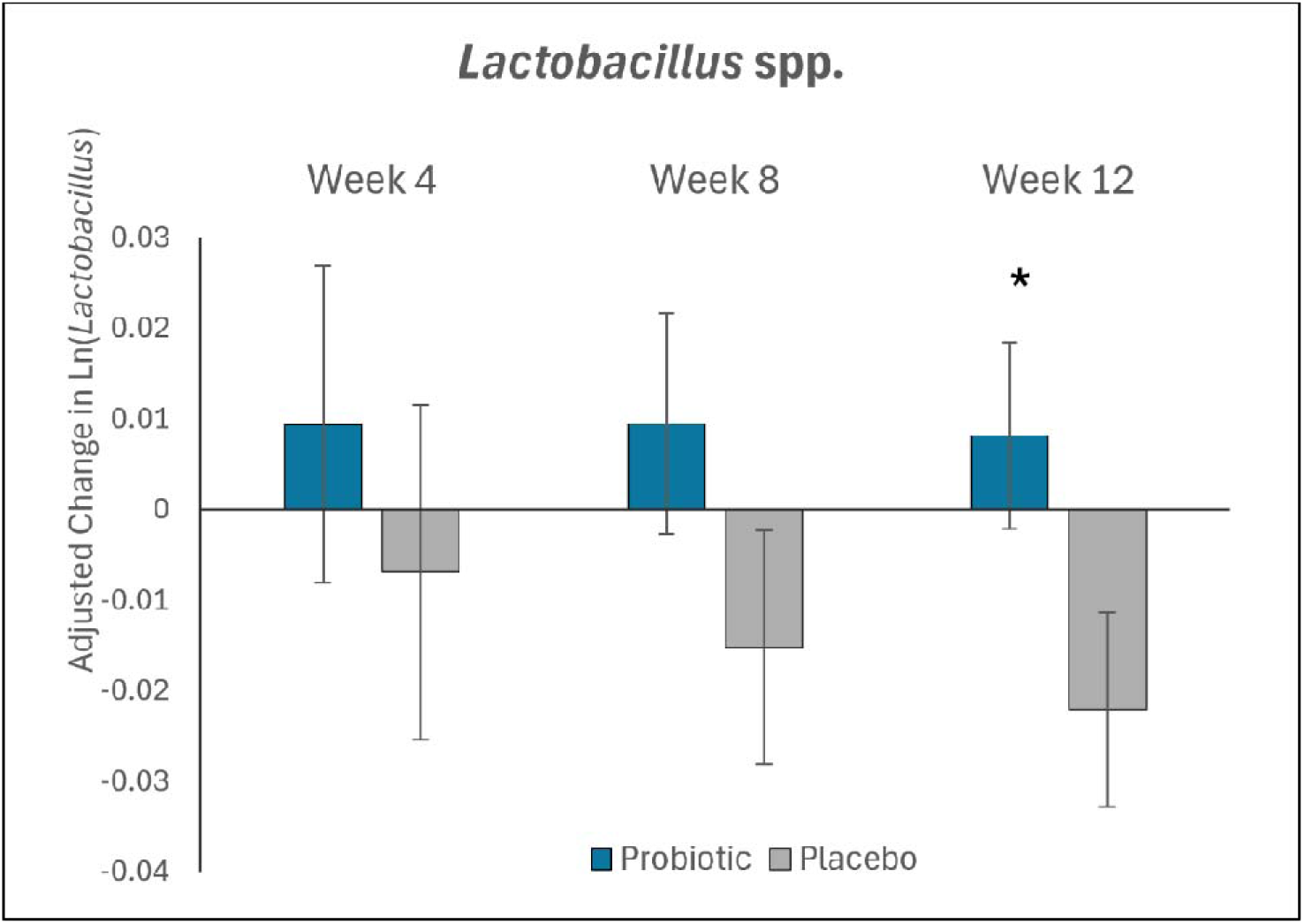
Changes from baseline in log-transformed *Lactiplantibacillus* abundance. Bars represent mean changes from baseline and their standard error. Asterisk (*) indicates significant difference vs. placebo (post-hoc analysis per timepoint from a Mixed Model for Repeated Measures, MMRM).

Salivary cortisol levels were also evaluated. The placebo group showed a 12% increase with respect to baseline concentration, while the probiotics group exhibited a 16% decrease. The between-group difference was not statistically significant.

## DISCUSSION

The results of the present study indicate that a specific strain of probiotic bacteria (*L. plantarum* KABP051) may be an effective supplement to enhance positive changes in body composition and reduced body weight, while also helping users to feel elevated energy (less fatigue) and improvements in other mood elements that may be helpful while they are embarking on a healthy lifestyle regimen, according to improvements in POMS questionnaires post-treatment.

The novelty of the current study is multi-faceted, including the healthy subject population (overweight or obese, but not diagnosed with any disease), supplementation with a unique strain of probiotic bacteria (*L. plantarum* KABP051), and elucidation of the relationship between microbiome balance, body composition, and mood.

Of particular interest is the fact that these subjects represent perhaps the largest subset of the American population who are not diagnosed with any disease or chronic condition, but yet routinely complain of symptoms related to weight gain (and difficulty with weight loss), high stress and fatigue, low mood and motivation, lack of enjoyment (anhedonia/burnout), and general malaise. Such individuals are poor candidates for prescription medications intended for severe metabolic conditions such as diabetes and obesity, or for psychological disease states such as major depression and anxiety disorders. Therefore, there is a pressing need for natural approaches to improve weight management and psychological mood states.

In the present study, body weight significantly improved with *L. plantarum* KABP051 supplementation. In comparison to placebo group, probiotic supplementation for 12 weeks was associated with a reduction in body weight of almost 2 kg (2.4% of body weight).

It is well-known that obesity increases the risk of multiple comorbidities, including cardiovascular disease, diabetes, hypertension, depression/anxiety, and several cancers, and carries an associated reduction in predicted lifespan.^47^ Even a modest loss of fat and total body weight of ∼3-5% has been shown to reduce morbidity and mortality risk, while improving mental well-being, provided that such losses can be sustained over time.^1,4^ Reductions in body weight of as little as 2-5% yet yield significant health benefits, such as reduction in cardiovascular risk factors, improved glycemic control, and reduced total testosterone and serum lipids in women affected by polycystic ovary syndrome.^47^

Similar studies support the effect of specific probiotic strains on weight reduction, though effects are known to be strain specific. A probiotic compound containing strains of *Bacillus subtilis* (LMG P-32899) and *Bacillus coagulans* (LMG P-32921) was supplemented to participants eligible for weight loss for 3 months. Results indicated the long-term use of at least double dose of this probiotic would be beneficial for weight loss than low doses; participants lost 0.6 kg in average with no observed weight changes between groups, while weight loss accounted for 1.4 kg on average after 6 months.^48^ Supplementing with resistant starch coupled with isoenergetic and balanced diets for 8 weeks helped reduce body weight 2.8 kg on average and improve insulin sensitivity, reshaping the microbiome structure and altered metabolites.^49^ *Bifidobacterium breve* BBr60 showed a significant reduction in body weight (4.67 kg) vs. placebo (2.82 kg) and body mass index after 12 weeks, helping regulate serum glucose, lipids, and liver function.^50^ Beyond body weight reduction, supplementation with *L. plantarum* KABP051 over 3 months resulted in an improvement of waist circumference and a reduction of 1.3% body fat. A combination of *Lacticaseibacillus paracasei* BEPC22 and *Lactiplantibacillus plantarum* BELP53, a human-derived probiotic, showed efficacy on the reduction of body fat by 0.58 % after supplementation during a similar period of time.^51^ Moreover, other recent clinical trials have shown that *Bifidobacterium lactis* B420, alone or in combination with prebiotic fibers, can be effective in controlling body fat mass, trunk fat mass, and waist circumference in overweight adults.^52–53^ In these studies, the relative change in body fat mass over a longer supplementation period of 6 months was approximately 3% in the per-protocol population with probiotic supplementation. Noteworthy, mood also improved after the intake of probiotic *L. plantarum* KABP051 in this study. Specifically, the POMS TMD global score, as well as the fatigue, confusion, and depression sub-scores decreased at the end of the intervention, with fatigue and confusion sub-scores showing significant differences compared to the placebo group. The POMS scores at baseline in our study were consistent with those observed in healthy populations. However, it is tempting to speculate that the intervention might lead to greater improvements in populations with specific mood disorders or higher stress levels, and future studies with *L. plantarum* KABP051 may explore the effects in such populations.

Emerging knowledge of the gut-brain axis provides a fundamental shift in both our understanding of and our ability to positively influence both metabolism (body composition) and psychology (mood) by directly modulating the microbiome.^3,5,7,9–11^ Microbiome modulation with targeted strains of probiotic bacteria represents a promising non-drug approach that enables people to simultaneously improve both their physical health and mental well-being. Previous studies have shown the effect of probiotic supplementation in reducing anxiety and stress. The consumption of *Lactiplantibacillus plantarum* DR7, a probiotic strain isolated from bovine milk, reduced DASS-42 total score in a population of moderately stressed adults and decreased serum cortisol levels.^5^ Interestingly, *Lactiplantibacillus plantarum* DR7 has high genetic and phenotypic similarity with *L. plantarum* KABP051.^54^ Likewise, in the present study, cortisol concentrations decreased in the group supplemented with probiotics compared to the placebo group; however, the difference was not statistically significant, likely due to the high variability in salivary cortisol measurements and the limited sample size. Further studies with larger cohorts are warranted to confirm the potential effects of *L. plantarum* KABP051 on mental well-being using biochemical biomarkers.

The potential benefits of microbiome modulation and gut-brain axis optimization for enhancing weight loss and/or improving mood may be confounded by the multifactorial nature of each condition, and by the disparate nature of the investigated interventions (e.g., single- or multi-strain formulations, supplementation dose, and duration of intervention). Nevertheless, recent scientific reviews have reported on several clinical trials showing an overall positive benefit of microbiome modulation for weight loss^44,48,51,55^ and mood improvement.^5,56–57^

Notably, this study purposely did not control for specific diet and exercise regimens, but the probiotic intervention was accompanied by weekly lifestyle education sessions on different topics. Therefore, our hypothesis is that the observed effects on body composition may result from improved mood, which could have enabled better self-regulation in maintaining a healthy lifestyle or avoiding unhealthy dietary habits. Additionally, our unpublished *in vitro* study using cultured enteroendocrine cells (STC-1 cells) showed that *L. plantarum* KABP051 induces glucagon-like peptide-1 (GLP-1) production, a hormone that plays a crucial role in regulating blood glucose levels, appetite, and digestion, which is related to appetite regulation. Thus, this raises the possibility that an effect on appetite regulation can also be at least partly responsible for the observed effects.

The weight loss and mood-enhancing effects of recent gut-brain axis interventions may be associated with their ability to alter both the structure and function of the microbiome, particularly the production of associated metabolites and signaling molecules including neurotransmitters, hormones/peptides, cytokines, short-chain fatty acids, etc.; remodeling of energy metabolism; modulation of gene expression related to appetite and thermogenesis; glucose homeostasis; lipid metabolism; and attenuation of HPA axis activity, among others.^44,58,59^ Unfortunately, the present study did not allow the measurement of many of those biomarkers, and therefore the mechanism underlying the effects of probiotic administration on body composition and psychological mood state through modulation of behavioral, mood and cognition responses requires further research.

Intervention with probiotic in the present study led to changes in microbiome composition. Specifically, *Lactobacillus* spp. increased significantly in the probiotic compared to the placebo group, in which this bacterial group decreased over the study period. Consequently, the mean difference in *Lactobacillus* spp. counts between both groups increased with time. The observed increase in *Lactobacillus* spp. counts is expected, given that the intervention consisted of a probiotic strain from the same genus. Furthermore, we hypothesize that changes in *Lactobacillus* spp. concentration may, at least in part, contribute to the observed beneficial effects on weight and body composition in our population. Studies have shown that probiotic supplements, most notably different strains of *Lactobacillus, Lactiplantibacillus*, and *Bifidobacterium* genus, have been shown to increase the number of beneficial bacteria (that produce anti-inflammatory signaling molecules such as SCFAs) and decrease the number of detrimental bacteria (that produce inflammatory signaling molecules such as LPS), ^60–62^ and thus may have beneficial effects in metabolism, promoting fat loss and elevating psychological mood state. Therefore, it is also worth noting that since participants felt better psychologically, those feelings of well-being may have contributed to high protocol adherence, and thus to the positive changes in body composition in a virtuous cycle of physical health and mental wellness supporting one another.

## CONCLUSIONS

Results from this randomized, double-blinded trial indicate that *L. plantarum* KABP051 may be an effective way to enhance weight loss, while also helping users improve their overall well-being with reduced fatigue and elevated energy levels that may be helpful when they are embarking on a healthy lifestyle program. Further investigations in larger populations are warranted to fully elucidate the underlying mechanisms by which *L. plantarum* KABP051 may exert their metabolic and psychobiotic effects.

## Data Availability

All data produced in the present work are contained in the manuscript

## ACKNOWLEDGEMENTS

The authors would like to thank the volunteers who contributed their samples, time, and focus to this study and Kaneka/AB-Biotics for providing the probiotic dietary supplements and funding the costs associated with microbiome assessments and psychological surveys.

## AUTHORS’ CONTRIBUTIONS

ST and AF designed the study protocol. JT coordinated the IRB submission, subject recruitment, and the study monitoring. BS and MO performed and oversaw microbiome assessments. All authors were involved in the preparation and presentation of these data.

## CONFLICTS OF INTEREST

ST and JT are employees of 3Waves Wellness, a contract research organization that was compensated for designing and conducting this trial. AF is an employee of Kaneka/AB-Biotics, the producer of the probiotic strain used in this study.

## ETHICAL APPROVAL

The study was approved by the Institutional Review Board (IRB) and Human Subjects Committee (HSC) at Marietta College (Marietta, OH, USA; Protocol #09nct162022-2) and registered at ClinicalTrials.gov (NCT06808061). This study was conducted following the guidelines laid down in the Declaration of Helsinki in 2013.

## Notes

### Clinical Trial

NCT06808061

### Funding Statement

Kaneka/AB-Biotics provided the probiotic dietary supplements and funded the costs associated with microbiome assessments and psychological surveys.

### Author Declarations

The Human Subjects Committee of Marietta College gave ethical approval for this work

